# The hidden burden of eye-care needs: how visual acuity definitions change estimates of need in 12 RAAB surveys across six countries

**DOI:** 10.64898/2026.09.19.26363481

**Authors:** Andrew Bastawrous, Marzieh Katibeh, Sergio Latorre-Arteaga, Michael Gichangi, Asad Khan, Shalinder Sabherwal, Moses Kasadhakawo, Sailesh Mishra, Rebecca Oenga, Furahini Godfrey Mndeme, Allen Foster

## Abstract

**Purpose:** Rapid Assessment of Avoidable Blindness (RAAB) surveys are widely used to estimate the prevalence and causes of blindness and vision impairment among adults aged 50 years and older. We examined how far estimates of eye-care need depend on whether presenting or pinhole acuity, and unilateral or bilateral impairment, is used.

**Methods:** We analysed 12 RAAB surveys undertaken between 2018 and 2023 in India, Nepal, Pakistan, Kenya, Tanzania and Uganda. Three person-level measures were compared: presenting visual acuity (PVA) in at least one eye, PVA in the better-seeing eye, and pinhole acuity in the better-seeing eye. Estimates were adjusted for the two-stage cluster design and age-standardised to the WHO world standard population.

**Results:** Of 48,185 participants examined (response rate 94.3%), vision impairment based on pinhole acuity in the better eye (9–19%) was substantially lower than on PVA in the better eye (18– 37%) or in at least one eye (33–51%). Increasing age (adjusted odds ratio [AOR] per year 1.106; 95% confidence interval [CI] 1.103–1.109), female sex (AOR 1.13; CI 1.08–1.19) and South Asian survey location (AOR 3.04; CI 2.78–3.33) were independently associated with impairment. Age-standardisation widened the difference between survey locations (prevalence ratio 1.50 crude, 1.66 standardised).

**Conclusions:** Substantial vision impairment persists in the surveyed districts of both regions. The large gap between presenting and pinhole acuity indicates a considerable addressable burden, and restricting estimates to bilateral impairment in the better-seeing eye omits a further substantial share of need. The higher prevalence in the South Asian locations most plausibly reflects survey site selection and uncorrected refractive error rather than a regional epidemiological difference.

## Introduction

Vision impairment and blindness remain important public health challenges worldwide despite the availability of effective interventions for many of their leading causes. At least 2.2 billion people are estimated to live with distance or near vision impairment globally, of whom at least one billion have impairment that could have been prevented or has yet to be addressed (1,2). Vision loss affects educational attainment, employment opportunities, productivity, independence, and quality of life, while also imposing substantial social and economic costs on individuals and societies (1,3).

The burden of vision impairment is disproportionately concentrated in low- and middle-income countries, where access to eye care services remains limited for many populations (1,4). Population ageing and demographic growth are expected to increase demand for eye care services, particularly in sub-Saharan Africa and South Asia, regions that continue to experience a high burden of avoidable vision impairment (2,4).

Robust epidemiological data are essential for understanding the magnitude and causes of vision impairment and for guiding eye health policy, planning, and resource allocation. RAAB surveys generate a broad set of indicators for this purpose, and fuller use of the information they already contain can sharpen estimates of met and unmet eye care need for service planning (1,2).

The Rapid Assessment of Avoidable Blindness (RAAB) methodology was developed to provide a practical, standardized, and cost-effective approach for generating representative population-based estimates of blindness and vision impairment among adults aged 50 years and older (5). Over the past two decades, RAAB has become the most widely used survey methodology for eye health planning globally. Its standardized sampling and examination protocols enable meaningful comparisons of vision impairment and its causes across different settings (5–7).

Each RAAB survey is designed to inform planning in the population it covers, and is reported on that basis. Because the protocol is standardised, records from separate surveys can also be brought together and analysed under common definitions, which is what makes direct comparison between settings possible. How vision impairment is defined, however, determines the magnitude of need that an estimate appears to describe. Presenting visual acuity in the better-seeing eye has conventionally been used for this purpose, aligning with WHO reporting categories and supporting international comparison (2). Presenting the three measures together for the same participants makes two quantities explicit. First, impairment affecting one eye only, which is set aside when attention is restricted to the better-seeing eye, even though it carries functional consequences and generates demand for services. Second, the share of impairment that refractive correction would resolve, given by the difference between presenting and pinhole acuity.

In this study, we pooled individual participant data from 12 RAAB surveys conducted in six countries in sub-Saharan Africa and South Asia. Our primary objective was to quantify how estimates of eye-care need change according to the visual acuity definition applied, comparing presenting acuity in at least one eye, presenting acuity in the better-seeing eye, and pinhole acuity in the better-seeing eye in the same participants, and to set these three measures side by side across countries. Secondary objectives were to describe the causes of blindness and vision impairment and the reported barriers to cataract surgery across these settings, and to examine the association of vision impairment with age, sex and region.

## Materials and Methods

### Study design and survey selection

This cross-sectional study analysed data from 12 Rapid Assessment of Avoidable Blindness (RAAB) surveys conducted in six countries in sub-Saharan Africa and South Asia between 2018 and 2023. The surveys were undertaken using standardized RAAB methodology to generate population-based estimates of blindness and vision impairment among adults aged 50 years and older. Countries with more than one survey contributed data from all eligible surveys, which were combined for country-level analyses. Details of the included surveys are presented in Table 1.

**Table 1.**
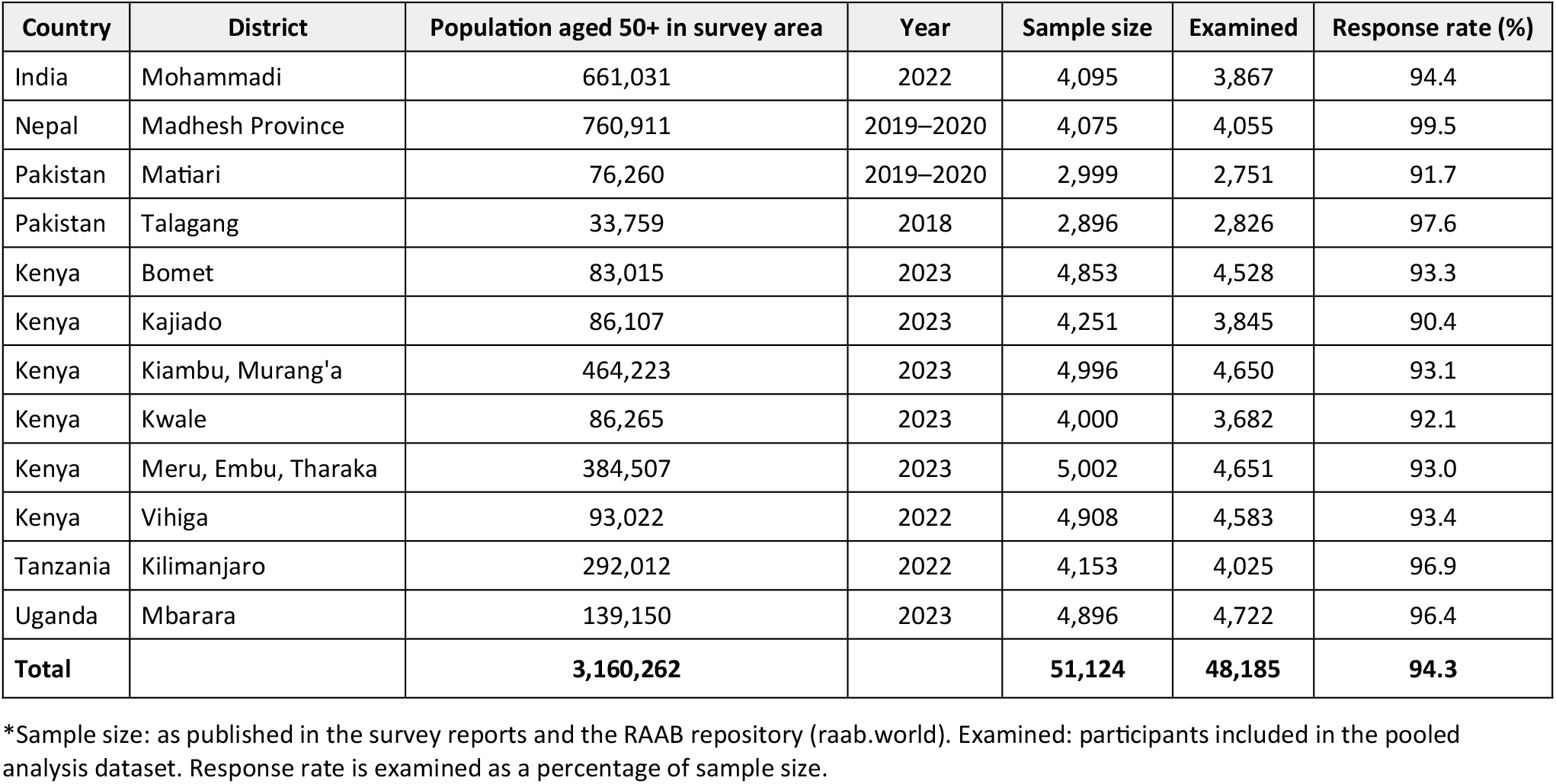
Source of data from 12 RAAB studies.

### RAAB survey methodology

Surveys were conducted using RAAB version 6 or version 7. Survey teams visited households to enumerate and examine eligible participants. Distance visual acuity was screened using Peek Acuity, the digital tumbling-E vision test integrated into the RAAB7 application (8), with printed single-optotype tumbling-E charts as the documented fallback where the digital test was unavailable. Both present the same optotype, and the digital test was developed and validated against paper-based tumbling-E charts. Participants with presenting visual acuity worse than 6/12 in either eye underwent pinhole testing and ophthalmic examination to determine the principal cause of vision impairment. Where the cause of vision impairment could not be established through the initial examination, additional assessment, including pupillary dilation when required, was performed.

Where more than one condition was present, the principal cause of vision impairment was assigned as the condition considered most readily treatable.

Across the 12 surveys, the sample size totalled 51,124 individuals aged 50 years and older, of whom 48,185 were examined and included in the pooled analysis dataset, giving an overall response rate of 94.3%.

### Definition of distance vision impairment

Distance vision impairment was classified according to World Health Organization definitions, based on presenting visual acuity in the better-seeing eye (2). Presenting visual acuity was measured with habitual correction, where available.

Mild vision impairment was defined as presenting visual acuity worse than 6/12 to 6/18. Moderate vision impairment was defined as worse than 6/18 to 6/60. Severe vision impairment was defined as worse than 6/60 to 3/60. Blindness was defined as presenting visual acuity worse than 3/60.

### Visual acuity metrics compared

Three person-level measures of distance vision impairment were derived for each participant and are reported throughout.

Presenting visual acuity in at least one eye classified a participant as impaired if either eye presented with acuity worse than 6/12, and therefore captured both unilateral and bilateral impairment.

Presenting visual acuity in the better-seeing eye classified a participant as impaired only if the better eye presented with acuity worse than 6/12, and therefore captured bilateral impairment alone. This is the conventional reporting measure and the basis for the WHO severity categories above.

Pinhole visual acuity in the better-seeing eye applied the same threshold after pinhole correction. Pinhole testing was performed in participants whose presenting acuity was worse than 6/12 in either eye, and provides a field-feasible approximation to best-corrected acuity.

Comparison of the first two measures quantifies the contribution of unilateral impairment. Comparison of the second and third quantifies the contribution of uncorrected refractive error.

### Barriers to cataract surgery

Participants were asked why cataract surgery had not been undertaken if they had visually impairing lens opacity in at least one eye, with presenting visual acuity worse than 6/18 in that eye that did not improve on pinhole testing. Up to two barriers were recorded per participant, in the order reported. This criterion identified 7,453 of the 48,185 participants examined.

### Statistical analysis

Analyses were conducted on individual participant records from all 12 surveys, restricted to the 48,185 participants who were examined. The survey design was declared with the 1,094 sampling clusters as primary sampling units. Prevalence estimates are presented as percentages with 95% confidence intervals derived from a linearised (Taylor series) variance estimator that accounts for clustering. Design effects ranged from 1.3 to 7.1 across countries and outcomes. Sampling weights were not applied, because RAAB’s combination of probability-proportional-to-size cluster selection with compact segment sampling is approximately self-weighting within a survey; a sensitivity analysis weighting each district by its 50 years and over population is reported for countries contributing more than one survey.

Prevalence was age-standardised by direct standardisation to the WHO world standard population, using the seven five-year age bands from 50–54 to 80 years and over, with standard weights rescaled to sum to one across the 50 years and over range. Standardised estimates were obtained as a linear combination of the design-based age-specific prevalences, so that confidence intervals account for both clustering and the covariance between age strata.

Associations between vision impairment in at least one eye and age, sex and region were examined using multivariable logistic regression, with age modelled as a continuous variable and region as a binary indicator contrasting South Asian with sub-Saharan African survey locations. Standard errors were linearised with clusters as primary sampling units. Cause-specific proportions are pooled across participants within each region so that larger surveys contribute proportionately, rather than averaged across countries.

All analyses were performed in Stata 14 (StataCorp, College Station, TX) using the svy suite of commands. The analysis code can be provided as Supporting Information.

### Ethical approval

Each RAAB survey received approval from the relevant national or institutional ethics review committee in the country where the survey was conducted, including India, Nepal, Kenya, Uganda, Pakistan, and Tanzania. Participants received information about the study in their local language and provided written informed consent before examination. All survey procedures followed the principles of the Declaration of Helsinki.

### Data security

Data storage, transfer, and access were governed by data-sharing agreements with local stakeholders and complied with applicable data protection requirements, including the European Union General Data Protection Regulation.

## Results

### Prevalence of distance vision impairment

A total of 48,185 participants aged 50 years and older were examined across 12 RAAB surveys conducted in six countries. Country-specific prevalence estimates of distance vision impairment (VI) are presented in Table 2. Figure 1 compares the prevalence of vision impairment based on presenting visual acuity (PVA) in at least one eye, PVA in the better-seeing eye, and pinhole visual acuity in the better-seeing eye.

**Table 2.** Prevalence of distance visual impairment in the 50+ population in 12 district-level RAAB surveys across 6 countries.

| Distance VI |  | India |  | Nepal |  | Pakistan |  | Kenya |  | Tanzania |  | Uganda |  |
| --- | --- | --- | --- | --- | --- | --- | --- | --- | --- | --- | --- | --- | --- |
|  |  | ≥1 eye | Better eye | ≥1 eye | Better eye | ≥1 eye | Better eye | ≥1 eye | Better eye | ≥1 eye | Better eye | ≥1 eye | Better eye |
| No VI | % | 48.8 | 70.3 | 51.0 | 67.5 | 50.9 | 62.7 | 67.2 | 81.5 | 65.0 | 77.9 | 67.4 | 81.9 |
|  | 95% CI | 46.8–50.7 | 68.6–72.0 | 48.5–53.4 | 65.1–69.9 | 47.4–54.4 | 59.5–66.0 | 66.3–68.1 | 80.8–82.2 | 62.3–67.7 | 75.9–80.0 | 65.8–69.0 | 80.6–83.1 |
| Mild VI | % | 10.8 | 8.8 | 14.3 | 15.3 | 18.8 | 21.1 | 7.5 | 6.7 | 10.3 | 9.4 | 8.3 | 5.9 |
|  | 95% CI | 9.6–11.9 | 7.8–9.8 | 13.0–15.6 | 13.9–16.7 | 16.1–21.5 | 18.1–24.0 | 7.2–7.9 | 6.3–7.0 | 9.2–11.4 | 8.2–10.6 | 7.5–9.2 | 5.2–6.6 |
| Moderate VI | % | 19.3 | 15.8 | 16.3 | 12.5 | 18.4 | 12.6 | 14.3 | 9.1 | 14.6 | 9.7 | 12.5 | 8.6 |
|  | 95% CI | 18.0–20.7 | 14.5–17.1 | 14.7–17.9 | 11.0–13.9 | 17.0–19.8 | 11.2–14.1 | 13.7–14.9 | 8.7–9.6 | 13.0–16.1 | 8.4–11.1 | 11.4–13.6 | 7.6–9.5 |
| Severe VI | % | 9.2 | 3.3 | 6.8 | 3.3 | 4.6 | 1.4 | 2.5 | 1.0 | 3.9 | 1.9 | 3.9 | 1.9 |
|  | 95% CI | 8.2–10.2 | 2.7–3.9 | 5.7–7.8 | 2.5–4.0 | 3.9–5.3 | 1.1–1.7 | 2.3–2.7 | 0.9–1.1 | 3.2–4.7 | 1.4–2.4 | 3.3–4.4 | 1.5–2.3 |
| Blind | % | 11.9 | 1.8 | 11.6 | 1.5 | 7.3 | 2.2 | 8.5 | 1.7 | 6.2 | 1.0 | 7.9 | 1.8 |
|  | 95% CI | 10.8–13.1 | 1.3–2.2 | 10.1–13.2 | 1.1–1.9 | 6.5–8.2 | 1.6–2.7 | 8.1–8.9 | 1.5–1.9 | 5.2–7.2 | 0.7–1.4 | 7.1–8.7 | 1.4–2.1 |
| Total VI | % | 51.2 | 29.7 | 49.0 | 32.5 | 49.1 | 37.3 | 32.8 | 18.5 | 35.0 | 22.1 | 32.6 | 18.1 |
|  | 95% CI | 49.3–53.2 | 28.0–31.4 | 46.6–51.5 | 30.1–34.9 | 45.6–52.6 | 34.0–40.5 | 31.9–33.7 | 17.8–19.2 | 32.3–37.7 | 20.0–24.1 | 31.0–34.2 | 16.9–19.4 |

≥1 eye: bilateral or unilateral vision impairment based on presenting visual acuity in at least one eye.
Better eye: bilateral vision impairment based on presenting visual acuity in the better-seeing eye.
95% confidence intervals account for the two-stage cluster design (1,094 clusters) using a linearised variance estimator.
CI: confidence interval.

**Figure 1.**
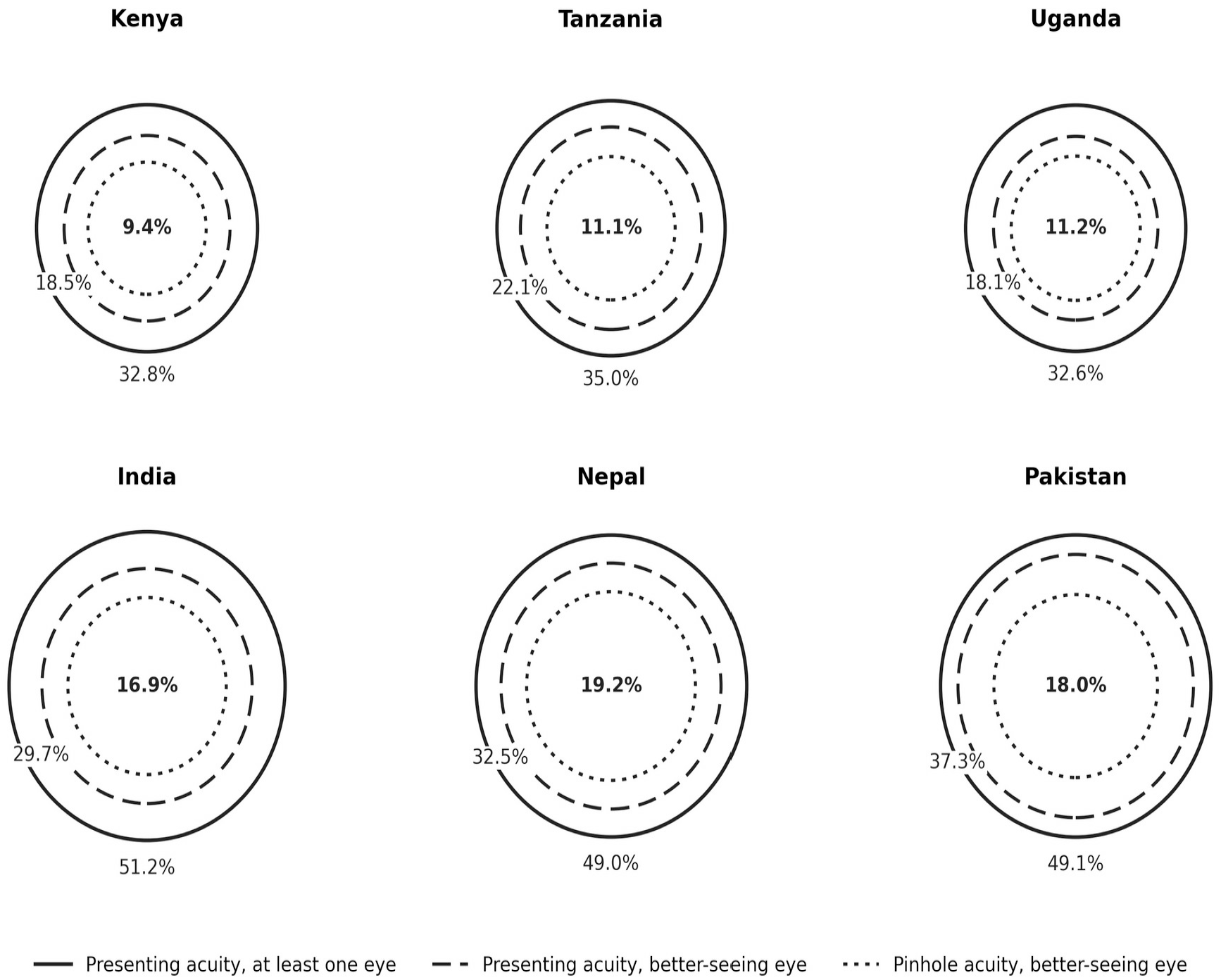
Prevalence of distance vision impairment (worse than 6/12) in adults aged 50 years and older, by visual acuity definition.

Substantial variation in the prevalence of vision impairment was observed across countries. Based on PVA in at least one eye, approximately half of adults aged 50 years and older in India (51.2%), Pakistan (49.1%), and Nepal (49.0%) had some degree of distance vision impairment, compared with 35.0% in Tanzania, 32.8% in Kenya, and 32.6% in Uganda.

Moderate vision impairment constituted the largest proportion of vision impairment in at least one eye in most countries, ranging from 12.5% in Uganda to 19.3% in India. Severe vision impairment was less common, ranging from 2.5% in Kenya to 9.2% in India, while blindness in at least one eye ranged from 6.2% in Tanzania to 12.0% in India. Mild vision impairment showed considerable variation, with the highest prevalence observed in Pakistan (18.8%) and the lowest in Kenya (7.5%).

When analysis was restricted to bilateral impairment, using PVA in the better-seeing eye, the estimated burden fell substantially. Bilateral vision impairment was highest in Pakistan (37.3%), followed by Nepal (32.5%) and India (29.7%), with lower prevalence in Tanzania (22.1%), Kenya (18.5%) and Uganda (18.1%). Bilateral blindness ranged from 1.0% in Tanzania to 2.2% in Pakistan, between three and eight times lower than the corresponding at-least-one-eye estimates, indicating that most blindness in these populations was unilateral.

### Potentially correctable vision impairment

Figure 1 demonstrates a substantial reduction in the prevalence of vision impairment when pinhole visual acuity was considered. The prevalence of vision impairment based on pinhole acuity in the better eye ranged from 9.4% in Kenya to 19.2% in Nepal, compared with 18–37% based on presenting visual acuity in the better-seeing eye and 33–51% based on presenting visual acuity in at least one eye (Table 3). The absolute difference between presenting and pinhole acuity in the better-seeing eye ranged from 6.9 to 11.0 percentage points in the African survey locations and from 12.8 to 19.3 percentage points in the South Asian locations. Because pinhole testing recovers less acuity than formal refraction, this difference is likely to understate the proportion of impairment that refractive correction could address.

**Table 3.** Prevalence of distance vision impairment (%) by visual acuity definition, with 95% confidence intervals adjusted for the cluster design.

| Country | Presenting, $\geq 1$ eye | Presenting, better eye | Pinhole, better eye |
| --- | --- | --- | --- |
| Kenya | 32.8 (31.9–33.7) | 18.5 (17.8–19.2) | 9.4 (8.9–9.9) |
| Tanzania | 35.0 (32.3–37.7) | 22.1 (20.0–24.1) | 11.1 (9.8–12.3) |
| Uganda | 32.6 (31.0–34.2) | 18.1 (16.9–19.4) | 11.2 (10.3–12.1) |
| India | 51.2 (49.3–53.2) | 29.7 (28.0–31.4) | 16.9 (15.5–18.2) |
| Nepal | 49.0 (46.6–51.5) | 32.5 (30.1–34.9) | 19.2 (17.1–21.2) |
| Pakistan | 49.1 (45.6–52.6) | 37.3 (34.0–40.5) | 18.0 (16.6–19.5) |

### Sex and age patterns

Figure 2a and 2b show the distribution of vision impairment by sex under each presenting acuity definition. Across most countries, females had a higher prevalence of vision impairment and blindness than males, both when assessed using vision in at least one eye and when restricted to bilateral vision impairment. The difference was most evident in India, Nepal, and Uganda, where females consistently exhibited lower proportions with no vision impairment and higher proportions with moderate vision impairment, severe vision impairment, or blindness.

**Figure 2.**
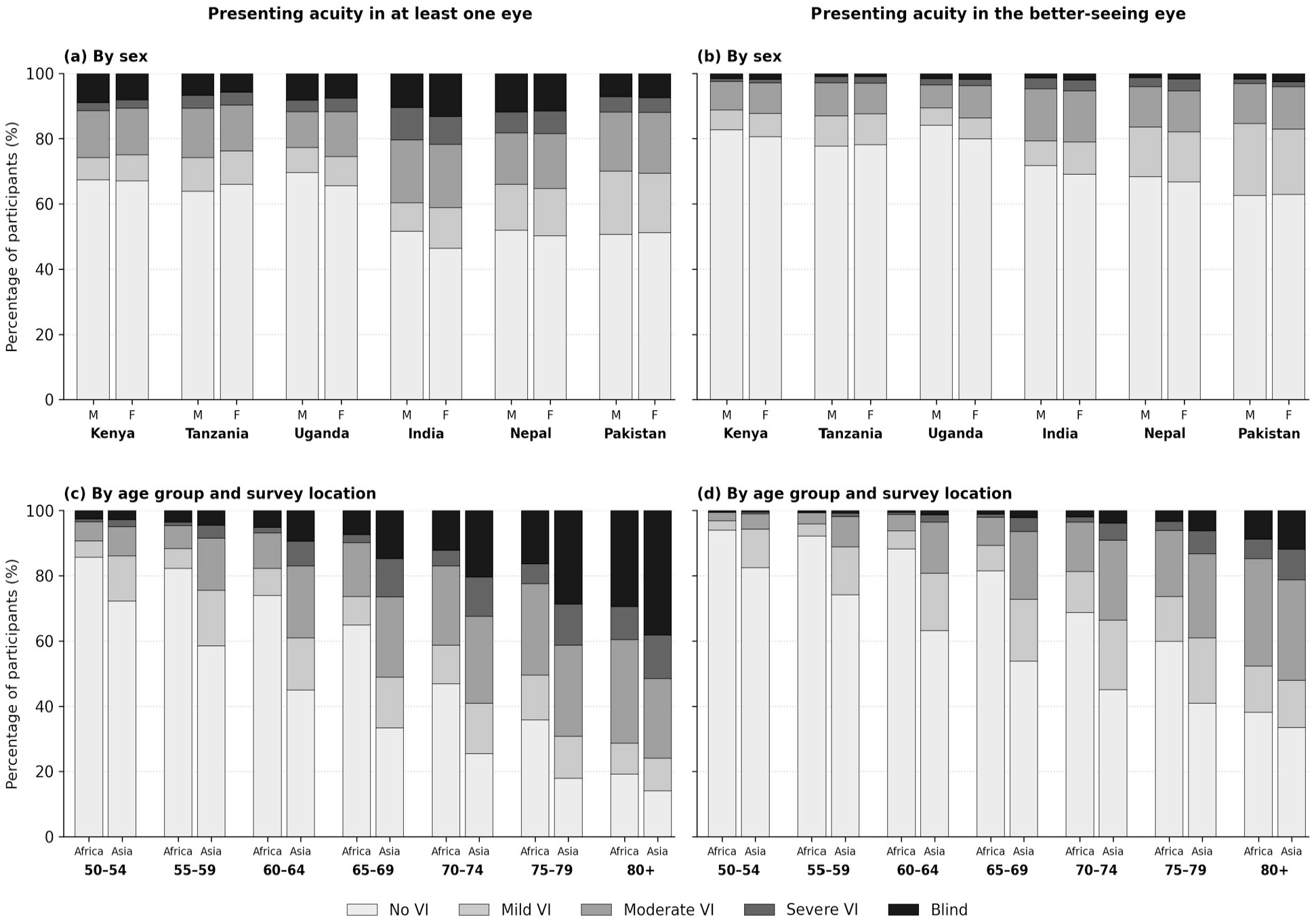
Distribution of distance vision impairment by sex, age and survey location. Left-hand panels (a, c) use presenting acuity in at least one eye; right-hand panels (b, d) use presenting acuity in the better-seeing eye. M: male; F: female. Africa: survey locations in Kenya, Tanzania and Uganda; Asia: survey locations in India, Nepal and Pakistan.

Figure 2c and 2d demonstrate a strong age-related increase in vision impairment in both groups of survey locations. The proportion of individuals without vision impairment declined progressively with age, while moderate vision impairment, severe vision impairment, and blindness became increasingly common. Across nearly all age groups, the burden of vision impairment appeared higher in South Asia than in sub-Saharan Africa, with the largest differences observed among older adults aged 70 years and above.

### Factors associated with vision impairment

Multivariable analysis demonstrated that the odds of vision impairment in at least one eye increased significantly with age (adjusted odds ratio [AOR] per additional year of age: 1.106; 95% confidence interval [CI]: 1.103–1.109). Females had higher odds of vision impairment than males (AOR 1.13; 95% CI: 1.08–1.19). Participants from South Asian survey locations had substantially higher odds of vision impairment than those from sub-Saharan African survey locations (AOR 3.04; 95% CI: 2.78–3.33). Because vision impairment was common in both groups, the odds ratio is considerably larger than the corresponding prevalence ratio: crude prevalence was 49.7% in the South Asian locations and 33.0% in the African locations, a prevalence ratio of 1.50 and a crude odds ratio of 2.00. Adjustment raises the odds ratio further because the South Asian samples were younger. The adjusted prevalence ratio was 1.80.

### Age-standardised prevalence

Because the surveyed populations differed in age structure, prevalence was also age-standardised to the WHO world standard population (Table 4). The South Asian samples were younger than the African ones, with a mean age of 60.3 compared with 62.9 years and 2.3–4.8% aged 80 years and over compared with 8.7– 9.4%. Standardisation therefore increased the estimate for the South Asian locations from 49.7% to 53.1% (95% CI 51.4–54.8) and slightly reduced the African estimate from 33.0% to 31.9% (95% CI 31.2–32.6), widening the prevalence ratio from 1.50 to 1.66. The difference between survey locations was therefore not attributable to differences in age composition.

**Table 4.**
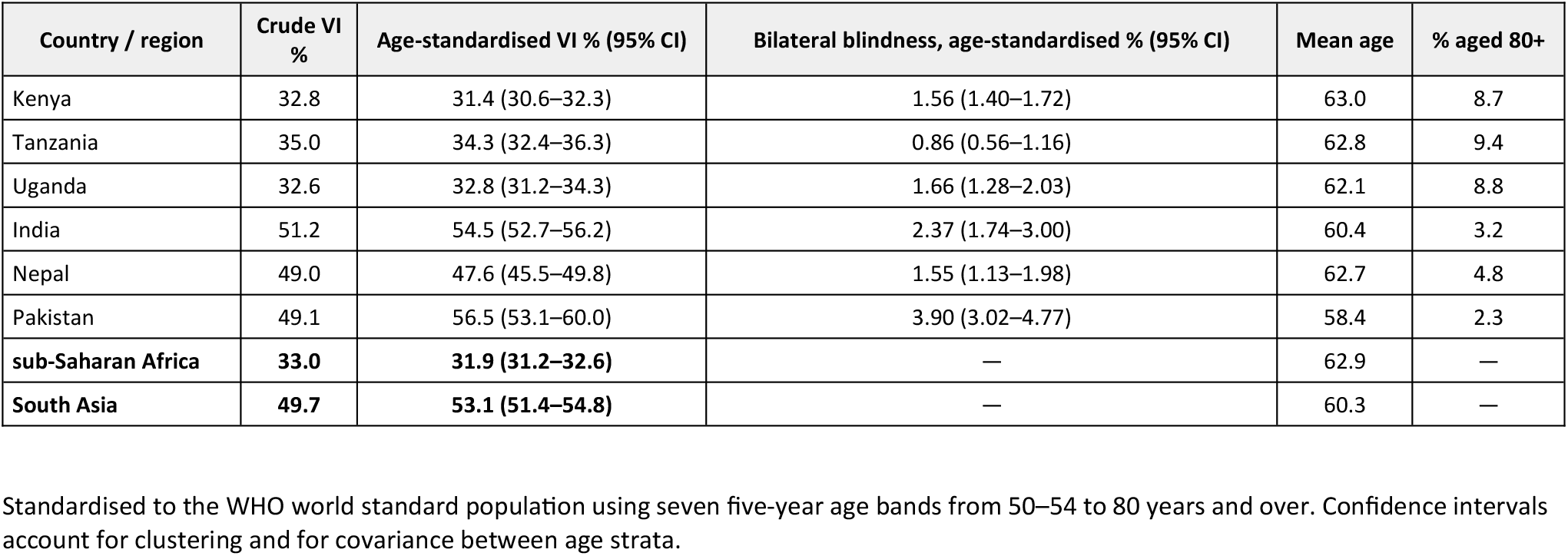
Crude and age-standardised prevalence of distance vision impairment in at least one eye, and age-standardised bilateral blindness, by survey location.

| Country / region | Crude VI % | Age-standardised VI % (95% CI) | Bilateral blindness, age-standardised % (95% CI) | Mean age | % aged 80+ |
| --- | --- | --- | --- | --- | --- |
| Kenya | 32.8 | 31.4 (30.6–32.3) | 1.56 (1.40–1.72) | 63.0 | 8.7 |
| Tanzania | 35.0 | 34.3 (32.4–36.3) | 0.86 (0.56–1.16) | 62.8 | 9.4 |
| Uganda | 32.6 | 32.8 (31.2–34.3) | 1.66 (1.28–2.03) | 62.1 | 8.8 |
| India | 51.2 | 54.5 (52.7–56.2) | 2.37 (1.74–3.00) | 60.4 | 3.2 |
| Nepal | 49.0 | 47.6 (45.5–49.8) | 1.55 (1.13–1.98) | 62.7 | 4.8 |
| Pakistan | 49.1 | 56.5 (53.1–60.0) | 3.90 (3.02–4.77) | 58.4 | 2.3 |
| sub-Saharan Africa | **33.0** | **31.9 (31.2–32.6)** | — | 62.9 | — |
| South Asia | **49.7** | **53.1 (51.4–54.8)** | — | 60.3 | — |

Standardised to the WHO world standard population using seven five-year age bands from 50–54 to 80 years and over. Confidence intervals account for clustering and for covariance between age strata.

The difference was present at every level of severity, with age-standardised prevalence ratios of 1.89 for mild, 1.39 for moderate and 2.71 for severe vision impairment in at least one eye, and 1.56 for blindness in at least one eye. Age-standardised bilateral blindness ranged from 0.86% (95% CI 0.56–1.16) in Tanzania to 3.90% (3.02–4.77) in Pakistan, with Nepal (1.55%) indistinguishable from Kenya (1.56%) and Uganda (1.66%).

In the sensitivity analysis weighting districts by their 50 years and over population, prevalence of vision impairment in at least one eye in Kenya, which contributed six surveys, was 31.1% compared with 32.8% unweighted; the corresponding figures for Pakistan, which contributed two, were 49.2% and 49.1%.

### Barriers to cataract surgery

Barriers to surgery were recorded for the 7,453 participants who had visually impairing lens opacity in at least one eye with presenting acuity worse than 6/18 that did not improve with pinhole testing. First-reported barriers varied substantially across the six countries (Figure 3a). “Need not felt” was the most frequent barrier in every country, ranging from 39.6% in Uganda to 81.5% in Nepal and 69.0% in India.

**Figure 3.**
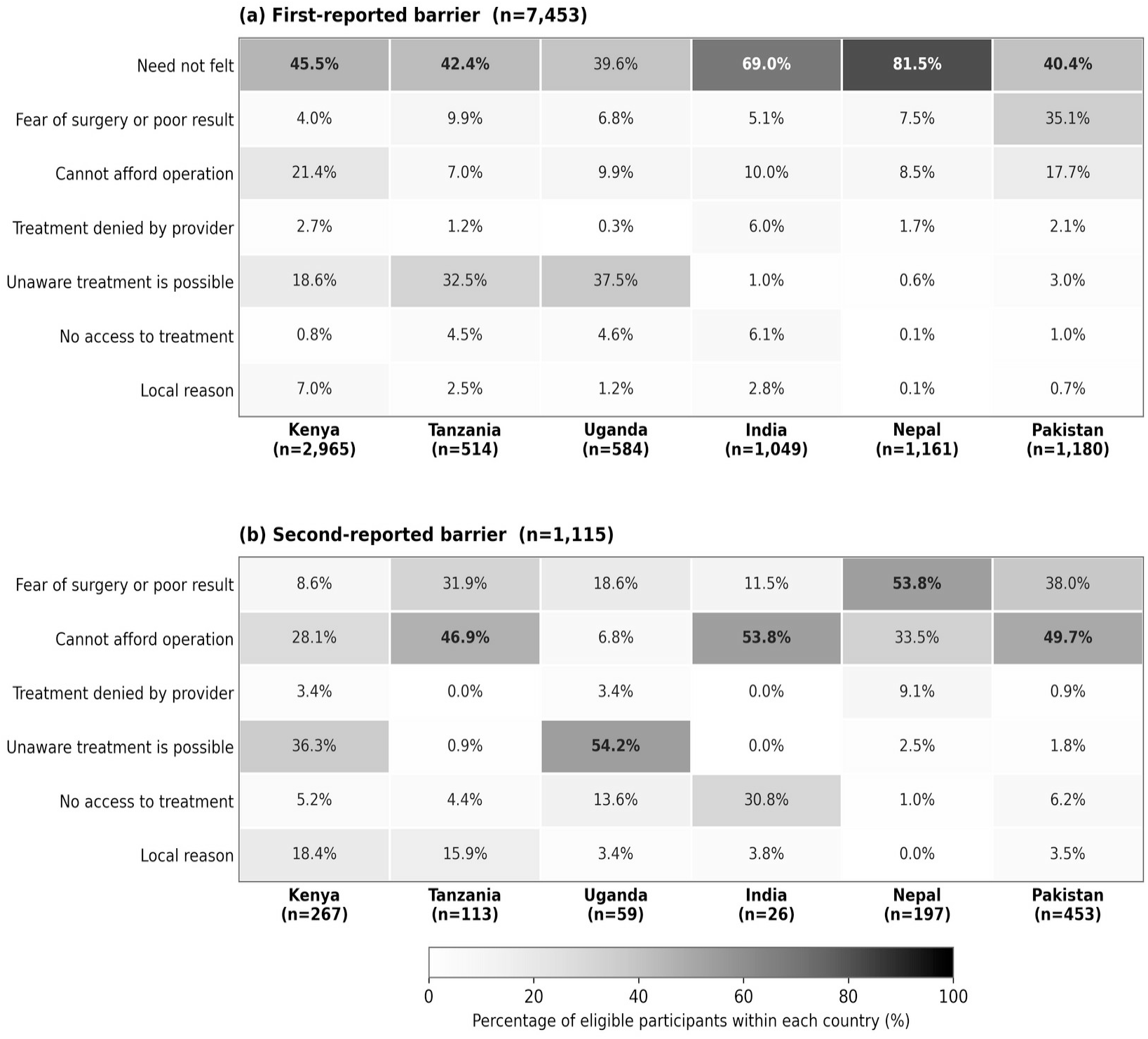
Reported barriers to cataract surgery among 7,453 participants with visually impairing lens opacity and presenting acuity worse than 6/18 not improving with pinhole in at least one eye: (a) first-reported barrier (n=7,453) and (b) second-reported barrier (n=1,115). Percentages are within country.

Beyond this, the profile differed markedly between settings. Unawareness that treatment is possible was prominent in the African locations, at 37.5% in Uganda, 32.5% in Tanzania and 18.6% in Kenya, but almost absent in the South Asian locations at 0.6% to 3.0%. Cost was the second most common barrier in Kenya (21.4%) and Pakistan (17.7%). Fear of surgery or of a poor result was reported by 35.1% of eligible participants in Pakistan, far above any other country (4.0% to 9.9%).

A second barrier was reported by 1,115 participants, 15.0% of those eligible, and its distribution differed from the first (Figure 3b). Cost became the leading second barrier in India (53.8%), Pakistan (49.7%) and Tanzania (46.9%), while fear of surgery predominated in Nepal (53.8%) and unawareness in Uganda (54.2%) and Kenya (36.3%). “Need not felt” did not appear as a second barrier, since participants giving it as their first response rarely offered another. Because only a minority reported a second barrier and some country denominators are small, 26 participants in India and 59 in Uganda, these proportions should be interpreted with caution. Taken together, the pattern indicates that multiple barriers operate concurrently, and that the predominant barrier differs by setting: awareness and cost in the African locations, cost and fear of surgery in the South Asian ones.

### Causes of blindness and vision impairment

Among 817 individuals with blindness in the better-seeing eye, untreated cataract was the predominant cause (57.4%), with marked differences between survey locations (Figure 4). In the African survey locations, cataract accounted for 51.5% of blindness while glaucoma contributed substantially at 19.8%, approximately seven times the South Asian figure (2.8%). In the South Asian locations, cataract dominated at 70.8%, followed by other corneal opacity at 14.0%, which was almost twice the African figure of 7.9%. Age-related macular degeneration and other posterior segment disease each accounted for a larger share of blindness in the African locations (5.1% and 5.3%) than in the South Asian ones (1.6% and 2.0%), whereas diabetic retinopathy was more prominent in South Asia (2.8% versus 0.9%). Country-specific data are presented in Supplementary Figures S1–S2.

**Figure 4.**
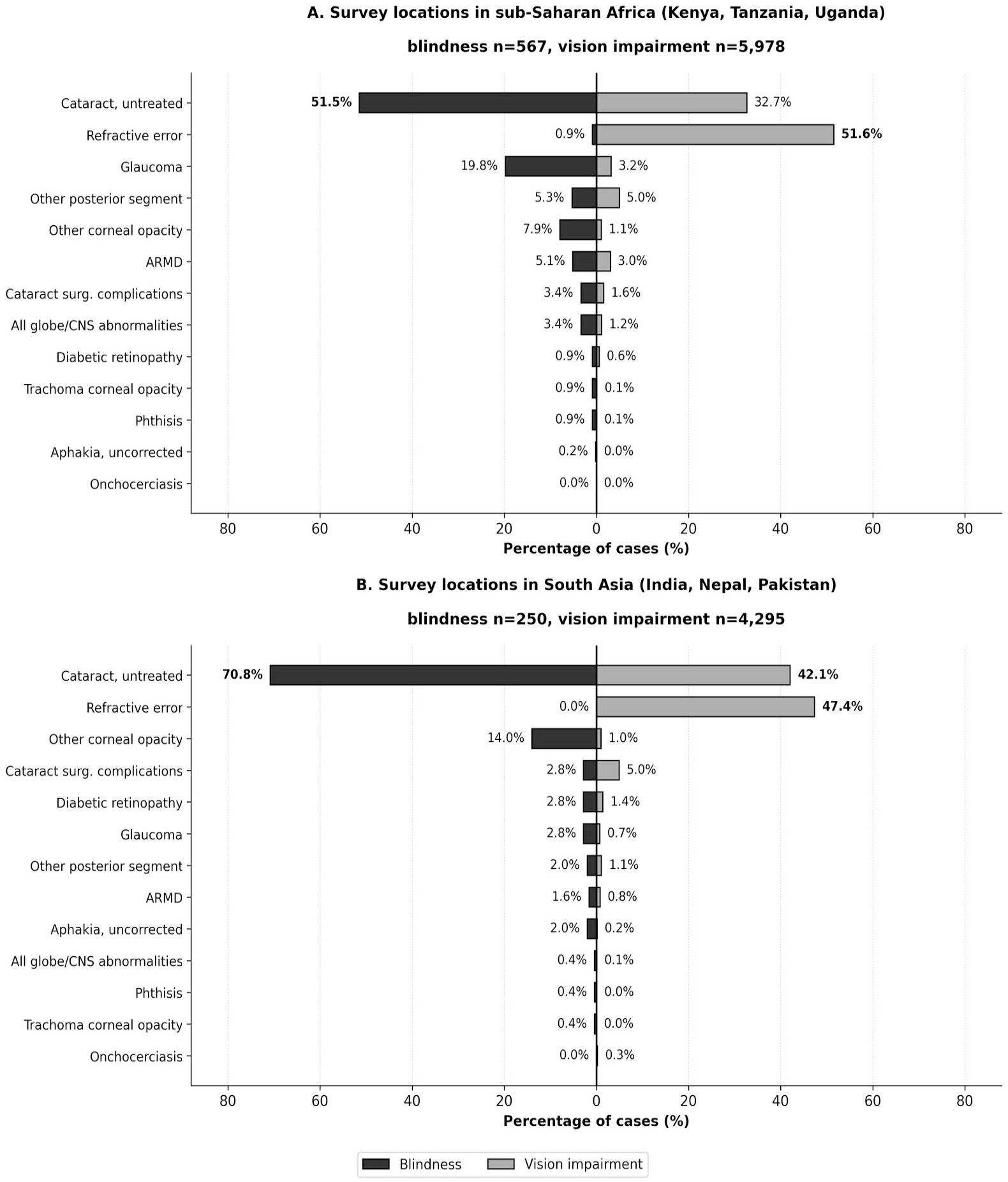
Causes of blindness and of vision impairment in the better-seeing eye, by survey location. Panel A: survey locations in sub-Saharan Africa. Panel B: survey locations in South Asia. In each panel, bars extending to the left show causes of blindness and bars extending to the right show causes of vision impairment. Percentages are of cases within each group of locations.

Among 10,273 individuals with mild to severe vision impairment in the better-seeing eye, refractive error dominated at 49.8%, reversing the blindness pattern. Variation between locations persisted: sub-Saharan Africa 51.6% versus South Asia 47.4% (with Pakistan at 53.5%). Untreated cataract remained substantial at 36.6% overall, with the African locations at 32.7% and the South Asian locations at 42.1%. Cataract surgical complications accounted for 5.0% of vision impairment in the South Asian locations compared with 1.6% in the African ones. Glaucoma, though rare overall (2.2%), was more than fourfold higher in sub-Saharan Africa (3.2%) than South Asia (0.7%). Other posterior segment disease and age-related macular degeneration were also more common causes in the African locations (5.0% and 3.0%) than in the South Asian ones (1.1% and 0.8%).

## Discussion

This analysis compared data from 12 RAAB surveys conducted across six countries in sub-Saharan Africa and South Asia and demonstrated substantial variation in the prevalence and severity of vision impairment.

Across all countries, vision impairment increased markedly with age and was more common among women than men. Survey locations in South Asia generally exhibited a higher burden of vision impairment than those in African ones. The prevalence estimates reported here are broadly consistent with previous population-based studies demonstrating that vision impairment remains a significant public health challenge in low- and middle-income countries despite substantial progress in eye care services globally (1,4,9). The strong age gradient observed in the present study mirrors findings from previous RAAB surveys and Global Burden of Disease analyses, which consistently identify increasing age as one of the strongest predictors of vision impairment and blindness (4,9,10). Similarly, the higher odds of vision impairment among women are consistent with longstanding evidence of gender disparities in access to eye care services and surgical treatment in many settings (1,4).

An important observation was the higher prevalence of vision impairment in the South Asian survey locations compared with the African ones. We interpret this as a difference between the specific populations surveyed rather than a regional epidemiological difference, for three reasons.

First, the surveys were conducted in districts selected for programme planning purposes rather than sampled to represent their countries or regions, so the contrast is between the populations surveyed and cannot be generalised. Examination protocols and visual acuity testing methods were comparable across sites, so differential measurement is unlikely to account for the difference, and the difference was present at every level of severity rather than concentrated near the 6/12 threshold where measurement error would be expected to act.

Second, differences in demographic structure and survival patterns may influence the composition of older populations included in surveys, particularly where mortality rates differ substantially (1,2). Global Burden of Disease analyses place the age-standardised prevalence of blindness in adults aged 50 years and over higher in eastern and western sub-Saharan Africa, at 40.0 and 42.2 cases per 1000 respectively, than in South Asia at 35.3 per 1000. (11) Our African survey locations produced substantially lower age-standardised bilateral blindness prevalence, between 0.86% and 1.66%, than these regional estimates would predict, whereas our South Asian locations, between 1.55% and 3.90%, were closer to or above them. This suggests that the African districts surveyed may be relatively well served, which is plausible given that all were established eye care programme areas at the time of survey. Establishing services is necessary but not sufficient; sustained reductions in blindness also depend on completing the referral pathway so that identified cases reach surgery.

Third, where our findings align closely with published regional estimates is uncorrected refractive error. Global Burden of Disease analyses report that uncorrected refractive error contributes a larger proportion of vision impairment in South Asia than in other regions, and that South Asia has the highest age-standardised prevalence of blindness due to uncorrected refractive error at 0.33%, compared with 0.11% in sub-Saharan Africa (12). In our data the absolute difference between presenting and pinhole acuity in the better-seeing eye was larger in the South Asian locations than in the African ones, consistent with a greater burden of uncorrected refractive error.

Taken together, the higher prevalence in the South Asian survey locations most plausibly reflects a combination of how survey districts were selected and a greater burden of uncorrected refractive error, rather than a general regional difference in eye health. The lower prevalence in the African survey locations should not be interpreted as evidence of better health system performance across sub-Saharan Africa, nor should the South Asian estimates be read as representative of that region.

A notable contribution of this study is the direct comparison of three acuity definitions applied to the same populations (Figure 1). Across all six countries, prevalence based on presenting acuity in at least one eye was approximately two to three times higher than prevalence based on pinhole acuity in the better-seeing eye, and the conventional RAAB measure — presenting acuity in the better-seeing eye — fell between the two. This finding highlights the large proportion of vision impairment that remains potentially addressable through refractive correction. While RAAB surveys traditionally emphasize estimates based on the better-seeing eye and often focus attention on blindness and bilateral vision impairment, health systems must respond to a broader spectrum of eye-care needs. Individuals with unilateral vision impairment, correctable refractive error, and less severe forms of visual loss may still experience important functional limitations and generate substantial demand for services (1,2,4). The difference between presenting and pinhole visual acuity therefore provides valuable information for planning comprehensive eye care programmes and estimating service requirements.

These findings also illustrate a broader challenge in population-based eye health surveillance. RAAB surveys have been instrumental in generating representative estimates of blindness and vision impairment among adults aged 50 years and older and remain one of the most important sources of eye health data globally (5–7). However, many eye-care needs occur outside the age groups traditionally included in RAAB surveys.

Conditions affecting children, working-age adults, and people with near vision impairment are not fully captured by conventional RAAB indicators, despite contributing substantially to the overall burden of eye conditions requiring care (1–3,13). Consequently, estimates of blindness and distance vision impairment among older adults should be viewed as an important component of population eye-care need rather than a complete description of service requirements. This distinction is becoming increasingly relevant as countries move towards integrated people-centred eye care approaches that seek to address eye health needs across the life course (1,14).

A companion analysis of routine programme data collected through community eye health programmes using the Peek platform in these same six countries addresses this gap directly by describing eye-care needs across the full age range and including near vision (15). The two studies are complementary rather than overlapping. The present analysis draws on probability samples of 48,185 adults aged 50 years and older and yields population prevalence estimates that are generalisable to the districts surveyed. The programme analysis describes 2,338,193 people of all ages who presented to screening services, and therefore characterises realised service demand rather than prevalence. The two datasets are distinct and no participant contributes to both. That designs with different sampling frames, age ranges and denominators independently identify substantial need beyond bilateral distance vision loss strengthens the inference that conventional survey indicators understate the scope of what eye care services are required to address.

That analysis reports higher overall eye care need in the sub-Saharan African programmes, the opposite direction to the contrast between survey locations observed here. The two are not directly comparable: the programme analysis covers all ages and a broader definition of need including near vision impairment and non-vision-impairing conditions. The higher prevalence of distance visual impairment in the South Asian locations most plausibly reflects survey site selection and uncorrected refractive error.

A prior comparison of RAAB data with programme reach in Talagang, Pakistan (16) illustrates how these survey estimates can be used to assess whether services are reaching the populations that need them, and the definitional distinctions described here directly affect the size of the denominator such assessments rely on.

Some of the constituent surveys have also been reported individually. Findings from the Mbarara survey have been published for Western Uganda, reporting prevalence, causes and coverage indicators for that population (17), and the diabetic retinopathy module of the Shahjahanpur and Lakhimpur Kheri survey has been reported separately for central Uttar Pradesh (18). The present study neither reproduces nor supersedes those reports. Its contribution is the harmonised pooling of 12 surveys under common definitions, and the comparison of three acuity definitions applied to the same participants, neither of which can be undertaken from single-survey reports.

The strengths of this study include the use of standardized RAAB methodology across multiple countries and the ability to compare findings across diverse settings using harmonized definitions and examination protocols. Prevalence was pooled across surveys within each country without weighting for district population size. In Kenya, which contributed six surveys, population-weighted prevalence was 31.1% compared with 32.8% unweighted, because the districts with the largest populations were not those contributing the largest samples.

However, some limitations should be considered. The included surveys were conducted in selected districts and regions rather than nationally representative samples, limiting the generalizability of country-level estimates. In addition, comparisons between survey locations presented here should be interpreted cautiously because the surveyed populations may differ in demographic composition, socioeconomic characteristics, and access to eye care services. Acuity after correction was assessed by pinhole testing rather than formal refraction, so the proportion of impairment attributable to uncorrected refractive error is likely to be underestimated. Near vision was not assessed, so the estimates presented describe distance vision needs only and understate total eye-care needs. Finally, the surveys spanned a six-year period including the COVID-19 pandemic, which may have affected service access and care-seeking differently across settings.

## Declarations

### Competing interests

AB: Co-founder — Peek Vision; MK and SL-A: Affiliation Peek Vision. No personal financial support related to this manuscript. The Peek Vision Foundation is a registered charity in the UK which wholly owns a not-for-profit company, Peek Vision Ltd. The Peek platform is designed as part of non-profit service delivery in resource-challenging settings in low- and middle-income countries, and is therefore developed for non-commercial interest. The remaining authors declared that this work was conducted in the absence of any commercial or financial relationships that could be construed as a potential conflict of interest.

### Funding

The authors received no specific funding for the present analysis. The RAAB surveys on which it draws were funded as follows. The surveys in Kenya, Tanzania, Uganda and Pakistan were funded by Christian Blind Mission (CBM). The survey in India was supported by Dr Shroff’s Charity Eye Hospital, New Delhi. The survey in Nepal was supported by Nepal Netra Jyoti Sangh, with technical support from the International Agency for the Prevention of Blindness. The funders had no role in the design of the present analysis, the preparation of this manuscript, or the decision to submit it for publication.

### Data availability

The data underlying this study are individual participant records from 12 Rapid Assessment of Avoidable Blindness surveys conducted by national eye care programmes and partner institutions in India, Nepal, Pakistan, Kenya, Tanzania and Uganda. Summary results for each survey are publicly available through the RAAB repository (raab.world). Data storage, transfer and access were governed by data-sharing agreements with local stakeholders in each country and complied with the European Union General Data Protection Regulation. De-identified aggregate data may be available from the corresponding author on reasonable request, subject to approval from the relevant national and international programme partners.

## Authors’ contributions

M.G., A.K., S.S., Mo.K., S.M., R.O. and F.G.M. carried out the studies and participated in data acquisition and interpretation. A.B. and S.L-A. participated in study design, data management and interpretation. A.B. and M.K. drafted the manuscript. M.K., A.B. and A.F. performed the statistical analysis and interpretation. All authors read and approved the final manuscript.

## Supplementary figures

**Supplementary Figure S1.**
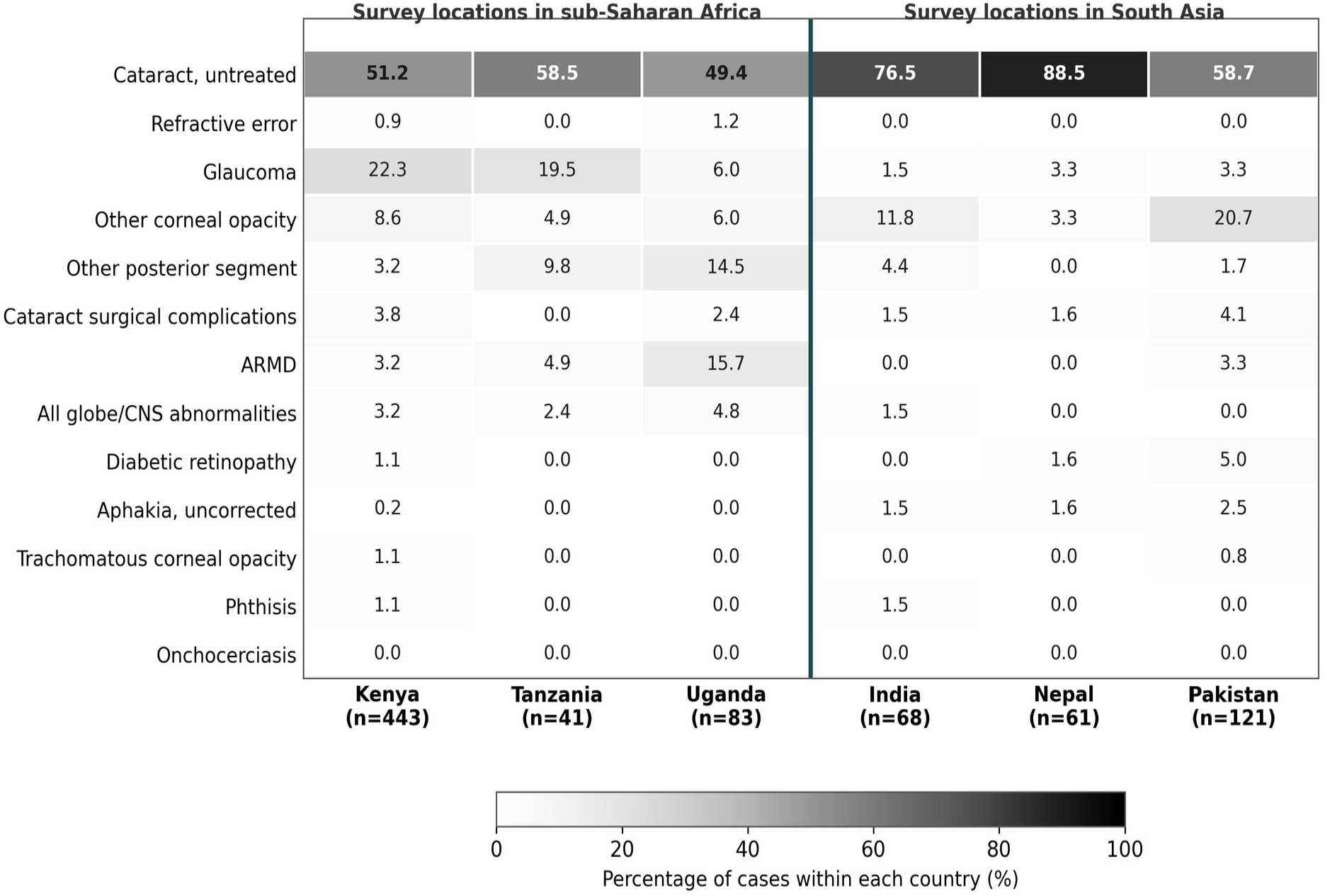
Causes of blindness in the better-seeing eye by country (n=817). Percentages are within country.

**Supplementary Figure S2.**
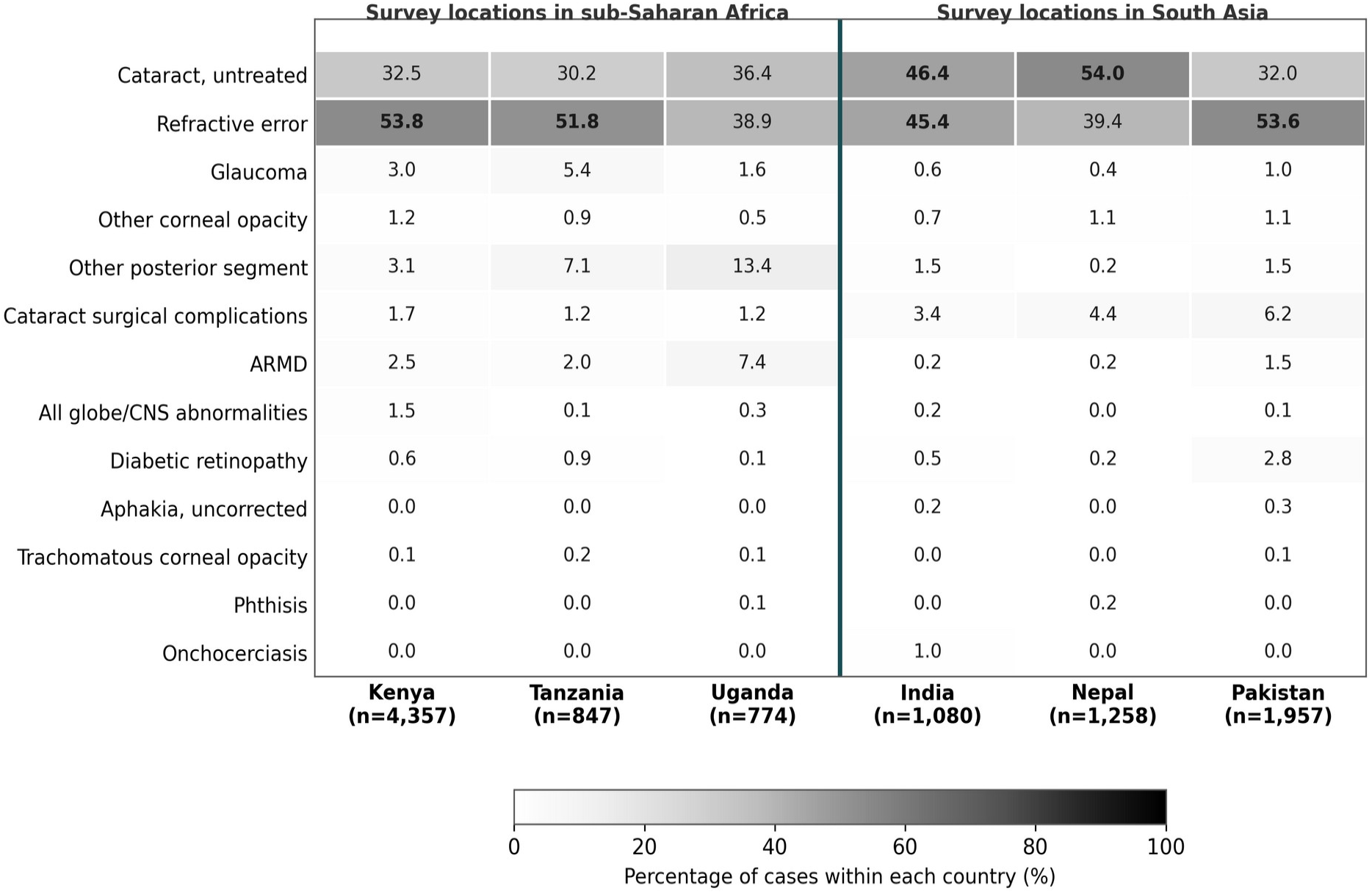
Causes of mild to severe vision impairment in the better-seeing eye by country (n=10,273). Percentages are within country.

## Notes

### Competing Interest Statement

The authors have declared no competing interest.

### Author Declarations

Each of the 12 Rapid Assessment of Avoidable Blindness surveys analysed in this study received ethical approval from the relevant national or institutional research ethics committee in the country where it was conducted, prior to data collection. In Nepal, approval was granted by the Nepal Health Research Council, Ministry of Health. Approval documentation for the remaining surveys is held by the respective national programme partners in India, Pakistan, Kenya, Tanzania and Uganda. All participants provided written informed consent before examination. The present study is a secondary analysis of de-identified individual participant records.

